# Using Exercise Intensity to Predict a Minimal Clinically Important Difference in the Six-Minute Walk Test in People with Chronic Stroke

**DOI:** 10.1101/2025.04.15.25325882

**Authors:** Kiersten M. McCartney, Pierce Boyne, Ryan T. Pohlig, Susanne M. Morton, Darcy S. Reisman

**Affiliations:** Department of Physical Therapy, University of Delaware, Newark, DE, USA; Biomechanics and Movement Science Program, University of Delaware, Newark, DE, USA; Department of Rehabilitation, Exercise and Nutrition Sciences, University of Cincinnati, Cincinnati, OH, USA; Biostatistics Core, Epidemiology Department, University of Delaware, Newark, DE, USA

## Abstract

**Background:** People with chronic stroke have significant impairments in their walking capacity. Minimal clinically important differences (MCIDs) can be used to interpret changes in patient outcomes following interventions. There is significant variability in the response to moderate-to-high walking interventions in people with chronic stroke. One reason for this response variability could be the lack of understanding of the threshold exercise dose needed to achieve an MCID. The purpose of this analysis was to determine the threshold of exercise training speed most predictive of a small (≥ 20m) or moderate (≥ 50m) clinically important difference in 6MWT in people with chronic stroke.

**Materials and Methods:** Participants with chronic stroke with a walking speed of 0.3-1.0m/s were randomized into a 12-week (1) fast-walking training or (2) fast-walking training and step-activity monitoring intervention. This analysis included participants (*n* = 129; age: 63.1 ± 12.5, 46% female) with complete pre- and post-intervention data. Exercise intensity was quantified as average training speed.

**Results:** Receiver operating characteristic curves analyzed whether training speed is predictive of attaining a clinically important difference in the 6MWT. Training speed had poor, non-significant accuracy of predicting a small (AUC [95% CI] = 0.584 [0.475 - 0.693], *p* = 0.131) or moderate (AUC [95% CI] = 0.597 [0.498 - 0.696], *p* = 0.056) change in 6MWT.

**Discussion:** The average walking training speed during this high-intensity walking intervention did not accurately predict which people with chronic stroke would attain a small or moderate clinically meaningful change in 6MWT distance.

## INTRODUCTION

In clinical practice, minimal clinically important differences (MCIDs) can be used to interpret changes in patient outcomes following interventions. MCIDs provide the smallest threshold value on a specific outcome (e.g., walking endurance) which indicate a patient-perceived beneficial change in that outcome was met.^1-3^ Meeting or exceeding an MCID is one factor clinicians can use to assess if the outcome of their intervention(s) were meaningful to their patient.

People with chronic stroke have significant impairments in their walking capacity and commonly cite improving walking as a priority for rehabilitation.^4,5^ MCIDs have been proposed for meaningful changes in walking endurance, measured by the six-minute walk test (6MWT), in people with stroke.^6-8^ There is no consensus on a singular MCID value for changes in 6MWT in those with stroke. However, this literature does provide a basis for reasonable MCID selection when trying to understand changes in 6MWT following intervention.

With this in mind, a recent meta-analysis examined changes in the 6MWT following moderate-to-high intensity walking exercise interventions, which have been shown to improve walking endurance in people with stroke.^9^ This analysis found high levels of variability in response to these walking interventions.^10^ It was estimated there is a 68% probability people with chronic stroke will have *at least a small clinically important difference* in walking endurance following moderate-to-vigorous intensity interventions, and only a 36% probability of having *a moderate clinically important difference* in walking endurance.^10^ One reason for this response variability could be the lack of understanding of the threshold exercise dose needed to achieve an MCID with these types of intervention.^11,12^

Cumulative evidence on exercise dose for walking training after stroke has demonstrated exercise intensity is a key ingredient to achieving greater changes in walking capacity.^9,13,14^ In most research interventions and the associated clinical translation of moderate-to-high walking interventions, training heart rate has been used as the measure of intensity.^9^ However, training speed, when considered as the metric of exercise intensity, is currently the most significant predictor of walking capacity outcomes in people with chronic stroke following high intensity interventions.^11,12^ It is unknown if there is a threshold training speed which would best predict an individual with stroke attaining a meaningful change in walking outcomes. Determining this threshold dose could assist clinicians by providing a more precise exercise prescription to reduce the exercise response variability.

The purpose of this analysis was to determine the threshold of exercise training speed that is most predictive of a small (≥ 20m) or moderate (≥ 50m) clinically important difference in 6MWT in people with chronic stroke.^7^ We hypothesized that higher average training speeds would be required to attain a moderate change compared to a small change in 6MWT distance following a fast-walking training intervention.

## METHODS

### Participants

This is a secondary analysis from a larger randomized controlled trial “*Promoting Recovery Optimization of Walking Activity in Stroke”* (“PROWALKS”; NIH1R01HD086362; NCT02835313). The full protocol and primary results have previously been published.^15,16^ Participants signed an informed consent prior to enrollment with all study procedures approved by the University of Delaware Institutional Review Board. The funders played no role in the design, conduct, or reporting of this study and the authors report there are no competing interests to declare.

Participants had their most recent stroke at least 6 months prior to enrollment, were 21-85 years old with self-selected walking speeds of 0.3-1.0m/s and could ambulate without the assistance of another person. Of the total 250 participants randomized to the PROWALKS clinical trial, this secondary analysis included the 169 participants randomized into the (1) fast-walking training (FAST, *n* = 89) or (2) fast-walking training and step activity monitoring combined intervention (FAST + SAM, *n* = 80).^17^ The 81 participants randomized to the step activity monitoring (SAM) intervention were excluded from this analysis as they did not undergo any high-intensity treadmill training.

### Clinical Evaluation

Prior to randomization, all participants underwent a baseline clinical evaluation which included collecting demographic and medical information. Demographic information included age and biological sex, and medical information included time since their most recent stroke (TSS), the Charlson Comorbidity Index (CCI), and the Activities-specific Balance Confidence Scale (ABC).^18-21^ The previously published protocol outlines all measures collected in the parent trial.^16^

Participants completed baseline measures of walking capacity, defined as what a person *can do* as assessed by standardized tests conducted in structured environments such as a clinic or laboratory.^22^ The six-minute walk test (6MWT) and 10-meter walk test (10mWT) are psychometrically strong measures and are recommended to quantify walking capacity in those with chronic stroke.^23,24^ The 6MWT may represent an individual’s walking endurance and can be influenced by their cardiovascular fitness and/or neuromotor function.^23,25-27^ The 10mWT can capture individuals self-selected (SSWS) and fastest walking speeds (FWS) over a short distance.^23,28^ Participants were instructed to “*walk at your normal pace*” (SSWS) or “*walk at your fastest pace*” (FWS) on the 10mWT and completed three trials at each speed, with the average of the trials recorded.^16^ The primary outcome of this analysis was the change in 6MWT distance.

### Exercise Intervention

Participants randomized to the FAST or FAST+SAM intervention received up to 36, 30-minute sessions of treadmill training over 12 weeks.^15,16^ The intervention protocol was designed to promote <u>continuous walking</u> at high (70-80% heart rate reserve) cardiovascular intensities.^16^

Training physical therapists were free to manipulate treadmill speed to achieve the target heart rate zone. All participants wore a safety harness which did not provide any body weight support. The exercise session was terminated if a participant’s response violated the guidelines set forth by the American College of Sports Medicine (ACSM) for individuals in phase III or IV cardiac rehabilitation.^29^ Heart rate was monitored continuously (Polar H10 chest-straps), with heart rate and treadmill speed recorded each minute.

### Exercise Intensity

For this analysis exercise intensity was quantified as the average training speed attained across the entire intervention. Values were averaged across every minute of training from the intervention. Treadmill speeds were converted from miles per hour to meters per second.

### Statistical Analyses

Receiver operating characteristic (ROC) curves are a graphical representation of whether a test or measure can accurately detect if an outcome of interest is attained (e.g., *a clinically meaningful change in walking distance*).^30,31^ If the area under the curve (AUC) of the ROC curve demonstrates appropriate prediction accuracy, Youden’s Index is a summary measure which identifies an optimal cutoff point by identifying the point at which sensitivity and specificity are maximized.^32,33^ This type of analysis could provide the threshold training speed required for people with chronic stroke to attain a meaningful change in 6MWT distance.

Two separate receiver operating characteristic (ROC) curves were used to analyze if walking training speed is predictive of individuals attaining a (1) small or (2) moderate change in the 6MWT.^34^ To complete these analyses, a binary outcome was used to classify the response of each individual as either “meeting” or “not meeting” the respective change in 6MWT. There is currently no consensus on the best approach to determining an MCID for medical interventions, including rehabilitation measures.^1,35^ Therefore, for this analysis, the MCID thresholds were chosen after reviewing the available literature on previously determined MCIDs for a change in 6MWT distance in people with stroke.^6-8^ To meet a small change in the 6MWT, participants had to have a change of at least 20m, and to meet a moderate change, participants had to have a change of at least 50m.^7^

The area under the curve (AUC) metric assessed the accuracy of each ROC curve, with AUC values of ≥ 0.9 indicating “excellent”, 0.8-0.9 indicating “good”, 0.7-0.8 indicating “fair/acceptable”, and < 0.7 indicating “poor” prediction accuracy, respectively.^34^ If an ROC curve had an AUC value of ≥ 0.7, indicating possible clinical utility,^36^ Youden’s Index was used to determine the threshold average training speed required to attain a small or moderate change in 6MWT.^32^ Youden’s Index is a summary measure of the ROC curve which identifies an optimal cutoff point for the variable of choice by identifying the point at which sensitivity and specificity are maximized.^32,33^

Using the same methodology, two additional ROC curves were used to analyze if walking training speed is predictive of individuals with chronic stroke attaining a (1) small or (2) moderate change in FWS (see Supplement).

## RESULTS

One-hundred twenty-nine participants randomized to the FAST (*n* = 68) or FAST+SAM (*n* = 61) intervention completed both baseline and post-intervention walking capacity measures. Participants were (mean±SD) 63±13 years old, 46% female, were 45±56 months (∼3.75 years) post-stroke (Table 1), and had on average a 44m increase in 6MWT distance (Table 2). Across the intervention, these participants attended 29 training sessions and walked for an average of 28 minutes per session at a training speed of 0.74 m/s (Table 3). There were no differences in baseline characteristics or training fidelity metrics between participants in the FAST or FAST+SAM interventions (Table 3, 4).

**Table 1:** Participant Characteristics.

| Characteristic | Participants (*n* = 129) |
| --- | --- |
| *Continuous* |  |
| Age (years) | 63.5 ± 12.5 |
| Time Since Initial Stroke (months) | 46.4 ± 57.8 |
| Body mass index (kg/m<sup>2</sup>) | 29.3 ± 6.3 |
| Self-selected walking speed (m/s) | 0.7 ± 0.2 |
| Fast walking speed (m/s) | 1.0 ± 0.3 |
| 6-Minute Walk Test (m) | 291.9 ± 117.2 |
| Average Baseline Steps/Day | 3,729.6 ± 1,862.6 |
| Activities Balance Confidence Scale | 74.0 ± 17.7 |
| Patient Health Questionnaire-9 | 3.7 ± 3.8 |
| Charlson Comorbidity Index | 3.3 ± 1.9 |
| *Categorical* |  |
| Sex | Female: *n* = 60 (46.5%) |
| Side of Hemiparesis | Left: *n* = 63 (48.8%)<br>Right: *n* = 60 (46.5%)<br>Bilateral: *n* = 6 (4.7%) |
| Assistive Device Use (yes/no) | Yes: *n* = 60 (46.5%) |
| Orthotic Device Use (yes/no) | Yes: *n* = 34 (26.4%) |
| Ethnicity | Hispanic or Latino: *n* = 3 (4.7%) |
| Race | Native American/American Indian/Alaska Native: *n* = 5 (3.9%)<br>Asian: *n* = 10 (7.8%)<br>Native Hawaiian or Pacific Islander: *n* = 0 (0%)<br>African American/Black: *n* = 33 (25.6%)<br>Caucasian/White: *n* = 86 (66.7%)<br>Prefer not to Respond: *n* = 1 (0.8%)<br>More than 1 Race: Yes (*n* = 8, 6.2%) |
| Beta-Blocker Medication | Yes: *n* = 49 (38.9%) |

*Continuous data presented as mean ± SD; Categorical data presented as n (% sample)*

**Table 2:** Changes in Walking Capacity Outcomes.

| **Outcome** | **ALL**<br>**(n = 129)** | **FAST**<br>**(n = 68)** | **FAST + SAM**<br>**(n = 61)** |
| --- | --- | --- | --- |
| **Six-Minute Walk Test (m)**<br>*Change from Baseline* | 44.0 ± 44.5<br>(-66.2 – 198.2) | 42.3 ± 46.8<br>(-60.7 – 198.2) | 46.0 ± 42.2<br>(-66.2 – 153.0) |
| **Fastest Walking Speed (m/s)**<br>*Change from Baseline* | 0.17 ± 0.17<br>(-0.13 – 0.73) | 0.16 ± 0.16<br>(-0.13 – 0.71) | 0.18 ± 0.17<br>(-0.12 – 0.73) |
| **Self-Selected Walking Speed (m/s)**<br>*Change from Baseline* | 0.13 ± 0.13<br>(-0.14 – 0.52) | 0.13 ± 0.13<br>(-0.11 – 0.48) | 0.13 ± 0.13<br>(-0.14 – 0.52) |

*Data presented as mean ± SD (range)*

**Table 3:** Exercise Dose Parameters.

| **Exercise Dose Parameter** | **ALL**<br>**(n=129)** | **FAST**<br>**(n = 68)** | **FAST + SAM**<br>**(n = 61)** |
| --- | --- | --- | --- |
| **Number of Sessions** | 29.0 ± 4.1<br>(19-36) | 26.1 ± 7.9<br>(21 – 36) | 29.2 ± 4.4<br>(19 – 36) |
| **Volume of Walking**<br>**(minutes)** | 823.0 ± 130.2<br>(427 – 1050) | 818.4 ± 124.9<br>(427 – 1050) | 828.2 ± 136.7<br>(431 – 1050) |
| **Average Heart Rate**<br>**Reserve (%)** | 63.6 ± 12.3<br>(19.6 – 123.3) | 64.3 ± 12.1<br>(45.9 – 123.3) | 62.7 ± 12.5<br>(19.6 – 120.7) |
| **Average Training Speed**<br>**(m/s)** | 0.74 ± 0.27<br>(0.11 – 1.79) | 0.74 ± 0.26<br>(0.23 – 1.38) | 0.73 ± 0.29<br>(0.11 – 1.79) |
| **Average Training Speed**<br>**(% of SSWS)** | 107.0 ± 32.0<br>(36.3 – 260.6) | 104.7 ± 26.1<br>(48.9 – 169.7) | 109.7 ± 37.5<br>(36.3 – 260.6) |

*Data presented as mean ± SD (range).*
*If a participant walked for all 30 minutes at each of a maximum of 36 sessions, the highest possible exercise volume was 1080 minutes.*

### Small Change in 6MWT Distance

Ninety-one (71.1%) participants had a change of at least 20m in 6MWT distance. Average training speed had poor, non-significant accuracy (AUC [95% CI] = 0.584 [0.475 - 0.693], *p* = 0.131) of predicting a small change in 6MWT (Figure 1).

**Figure 1.**
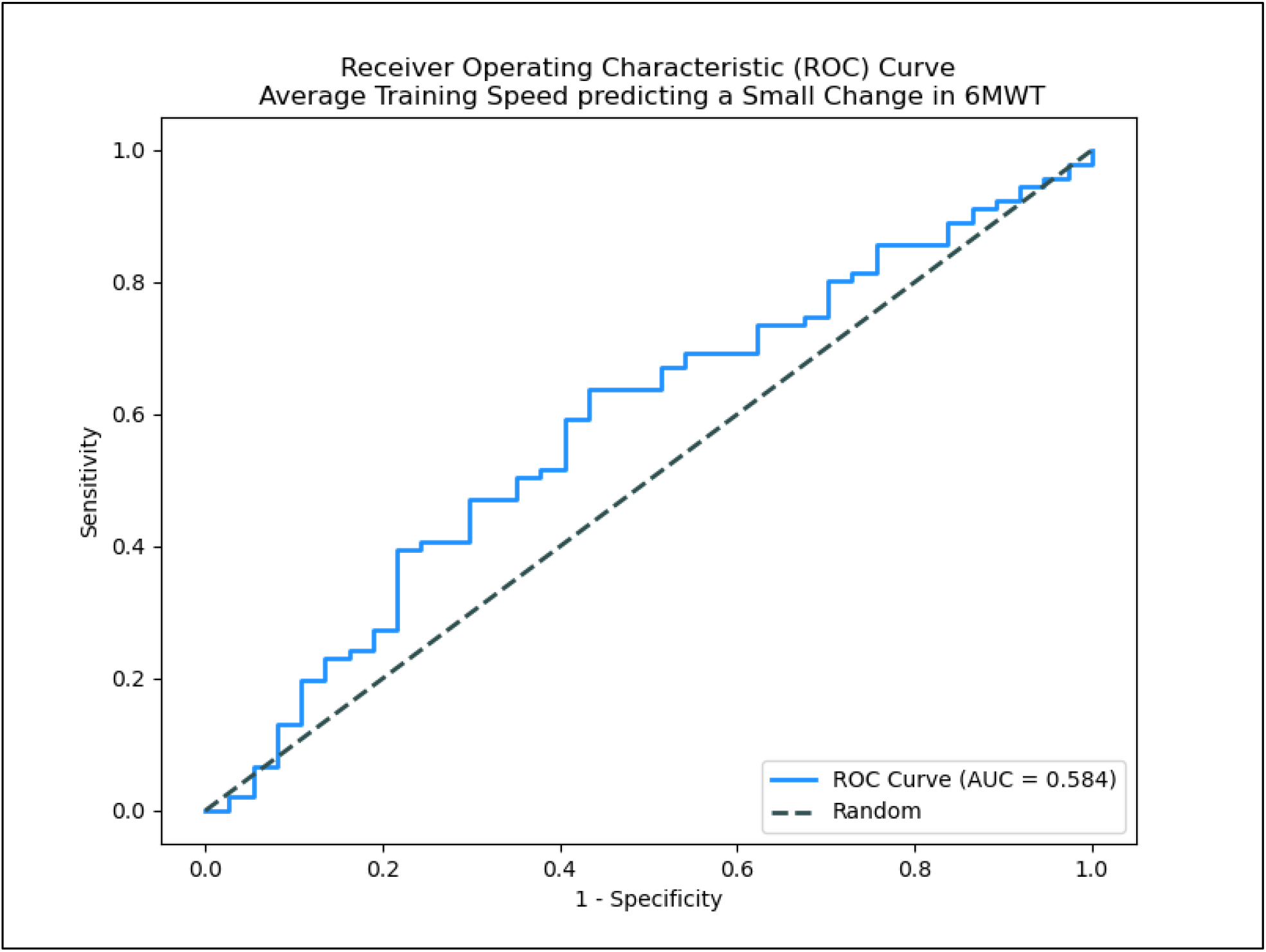
*6MWT = Six Minute Walk Test; small change = a pre-to-post change in 6MWT distance of ≥ 20 meters*.

### Moderate Change in 6MWT Distance

Forty-nine (38.3%) participants had a change of at least 50m in 6MWT distance. Average training speed had poor, non-significant accuracy (AUC [95% CI] = 0.597 [0.498 - 0.696], *p* = 0.056) of predicting a moderate change in 6MWT (Figure 2).

**Figure 2.**
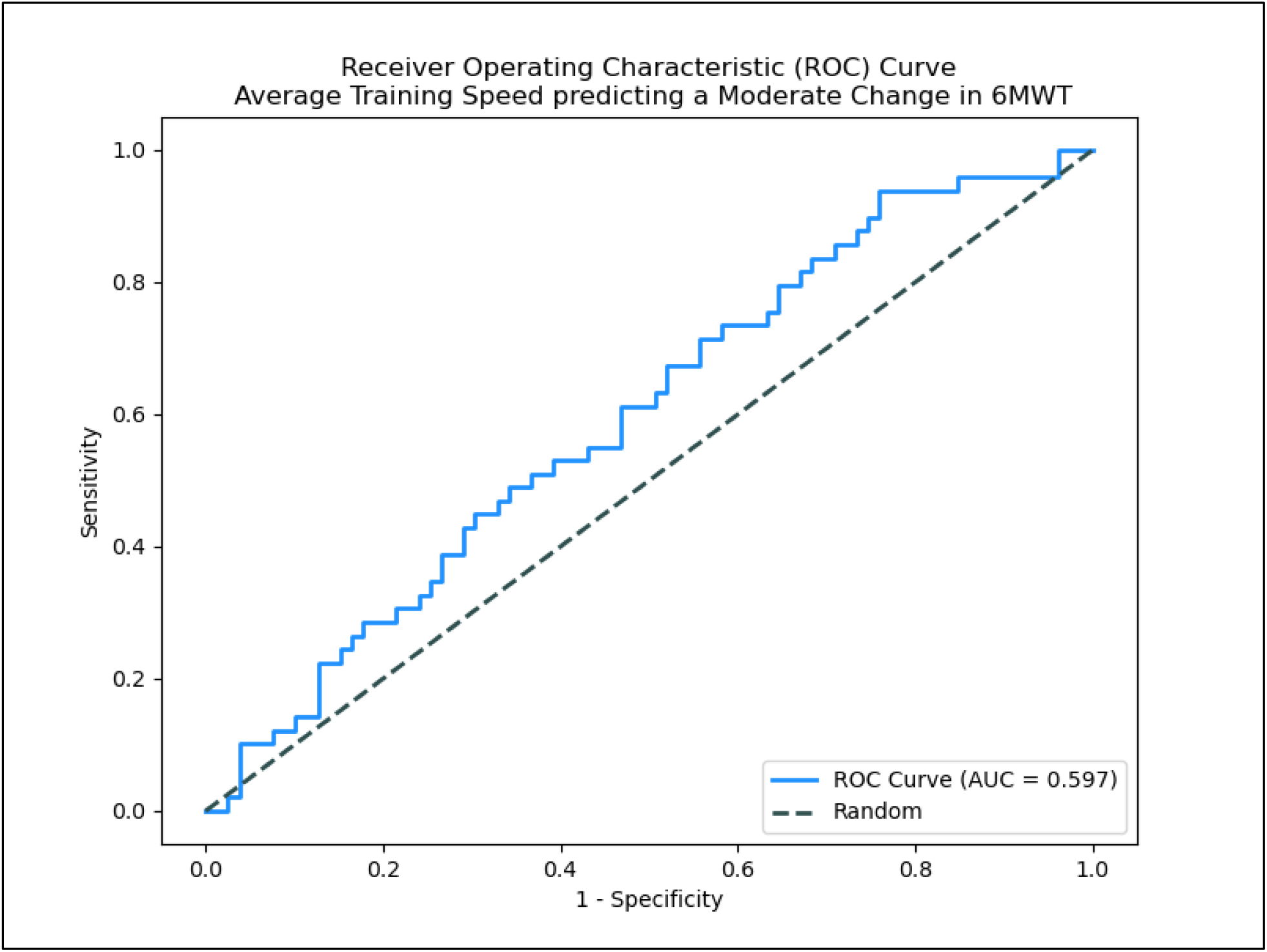
*6MWT = Six Minute Walk Test; moderate change = a pre-to-post change in 6MWT distance of ≥ 50 meters*.

## DISCUSSION

The average walking training speed during this high intensity walking intervention did not accurately predict which people with chronic stroke would attain a small or moderate clinically meaningful change in 6MWT distance. Since individuals with chronic stroke prioritize walking recovery in rehabilitation,^9,37-41^ and only 38.7% of American physical therapists feel confident prescribing aerobic exercise for this group,^42^ this analysis was an important step toward developing a precise exercise prescription to improve walking capacity in people with chronic stroke.

Previous studies of walking interventions in people with stroke have explored (1) what parameters of exercise dose are key or “active” ingredients in walking interventions,^43-49^ and most recently, (2) what metric of exercise intensity is the strongest predictor to achieving changes in walking capacity outcomes.^9,11,12^ Collectively the results of this research have determined that intensity is a key ingredient for walking interventions, and training speed is a significant predictor of changes in walking endurance in people with chronic stroke.^11,12^ A next step in optimizing exercise dose for clinical application is to decipher the therapeutic dose required to achieve clinically meaningful changes, which was the motivation for this analysis. However, the results found here demonstrate that training speed alone was not sufficient to identify a specific threshold exercise dose needed to achieve meaningful changes in walking capacity outcomes in people with chronic stroke.

Training speed during a walking intervention is only one parameter of overall exercise dose. Exercise dose includes multiple parameters including the intervention duration, duration of each session, frequency of sessions, and intensity of training within each session, making dosing within an exercise intervention highly complex.^35,36^ There is a confluence of evidence pointing to intensity being a key factor in people with chronic stroke achieving changes in their walking capacity following moderate-to-high intensity walking intervention.^9,12,13^ However, the current results suggest one exercise dose parameter may not be sufficient to predict a dichotomized response of a small or moderate change in a walking capacity outcome. Future studies could attempt to determine an exercise dose metric which incorporates multiple parameters. Prior work in our lab, however, has demonstrated how coupling parameters of exercise dose can lead to ambiguity on how the parameters are interacting with one another (McCartney et al., 2025, in press *JNPT*). Coupling parameters of exercise dose can reduce the ability to provide guidelines on what dose is occurring. This is because the combined arbitrary dosing metric could be due to a high volume/low intensity combination or a low volume/high intensity. Further work is needed to determine if there is a metric with improved prediction accuracy to determine which individuals with chronic stroke will meet an MCID.

While minimal clinically important differences (MCID) can be used to interpret whether a patient achieved a “meaningful change”, it is important to consider (1) the method used to calculate the MCID, and (2) the specific characteristics of the population used to calculate the MCID.^1,35,50,51^ As there is no consensus on the best approach to determining an MCID for rehabilitation measures,^1,35^ the previously defined MCID thresholds for a change in 6MWT distance in people with stroke were thoroughly assessed and are briefly reviewed here.^6-8^

Perera et al., used 3 different methods to determine initial estimates for a small or moderate change in the 6MWT in 100 individuals with stroke.^7^ When assessing the results across these methods, the recommended criterion was 20m for a small meaningful change and 50m for a substantial meaningful change.^7^ Tang et al., used a correlation analysis to assess the relationship between the measured change in 6MWT distance and perceived change in walking ability in 22 individuals with chronic stroke.^6^ The difference in 6MWT change between the groups was 34.4m [95% CI = 17.2-51.6].^7^ While the Tang et al. study had a much smaller sample size, and less rigorous analytical techniques, the 95% confidence interval is very similar to the recommended MCIDs from Perera et al. Lastly, Fulk & He used two-different anchors to determine MCID values for the 6MWT.^8^ However, the ROC curve analysis for both anchors (modified Rankin Scale, Stroke Impact Scale) had poor prediction accuracy (AUC = 0.66, 0.59, respectively).^8^

Previous literature has demonstrated how MCIDs can significantly differ based on the analysis used to derive it.^50^ Using 17-unique analyses, Franceschini et al., found extreme heterogeneity in the MCIDs calculated, including up to a 14-time difference in the threshold value determined between analyses.^50^ This calls into question if MCIDs can reliably account for a “meaningful” change. Furthermore, dichotomizing a previously continuous outcome (*such as a change in 6MWT*) can lead to a substantial loss of information.^52^ While this is an unavoidable limitation of ROC analyses, this loss of information in combination with the current uncertainty of MCID thresholds for the 6MWT in people with chronic stroke is likely contributing to the inability to determine the threshold training speed required to accurately predict a small or moderate meaningful change.

## Conclusions

While ROC curves have been useful in other areas of medicine to determine thresholds for diagnosis, exercise training speed had unreliable accuracy for predicting a small or moderate change in 6MWT in people with chronic stroke. There is current ambiguity in determining MCID thresholds for rehabilitation outcomes and uncertainty about whether these thresholds accurately assess a meaningful change in walking endurance in people with chronic stroke. Collectively, these analytical and methodological considerations, paired with the complexity of exercise dosing, make it difficult to determine the threshold exercise dose required to meet a small or moderate change in 6MWT in people with chronic stroke. This area of study is worthy of continued exploration as elucidating the threshold exercise dose required to achieve a meaningful change in walking capacity has the potential to reduce response variability to walking interventions and improve rehabilitation outcomes for people with chronic stroke.

## Data Availability

Deidentified data and relevant documents from the parent clinical trial are available via the NICHD DASH repository (https://dash.nichd.nih.gov/) upon reasonable request approved by the repository.

https://dash.nichd.nih.gov/study/425019

Appendix

### Additional Analyses

Two separate receiver operating characteristic (ROC) curves were used to analyze if exercise training speed is predictive of individuals attaining a (1) small or (2) moderate change in the FWS.^34^ A small FWS change was defined as a change of 0.1m/s with a moderate change defined as a change of 0.2m/s.

### Results

#### Small Change in FWS

Seventy-nine (61.2%) participants had a change of at least 0.1m/s in FWS. Average training speed had poor, significant accuracy (AUC [95% CI] = 0.671 [0.577 - 0.765], *p* < 0.001) of predicting a small change in FWS (Supplementary Figure 1).

#### Moderate Change in FWS

Forty-nine (38.3%) participants had a change of at least 0.2m/s in FWS. Average training speed had poor, significant accuracy (AUC [95% CI] = 0.697 [0.605-0.790], *p* < 0.001) of predicting a moderate change in FWS (Supplementary Figure 2).

**Supplemental Figure 1.**
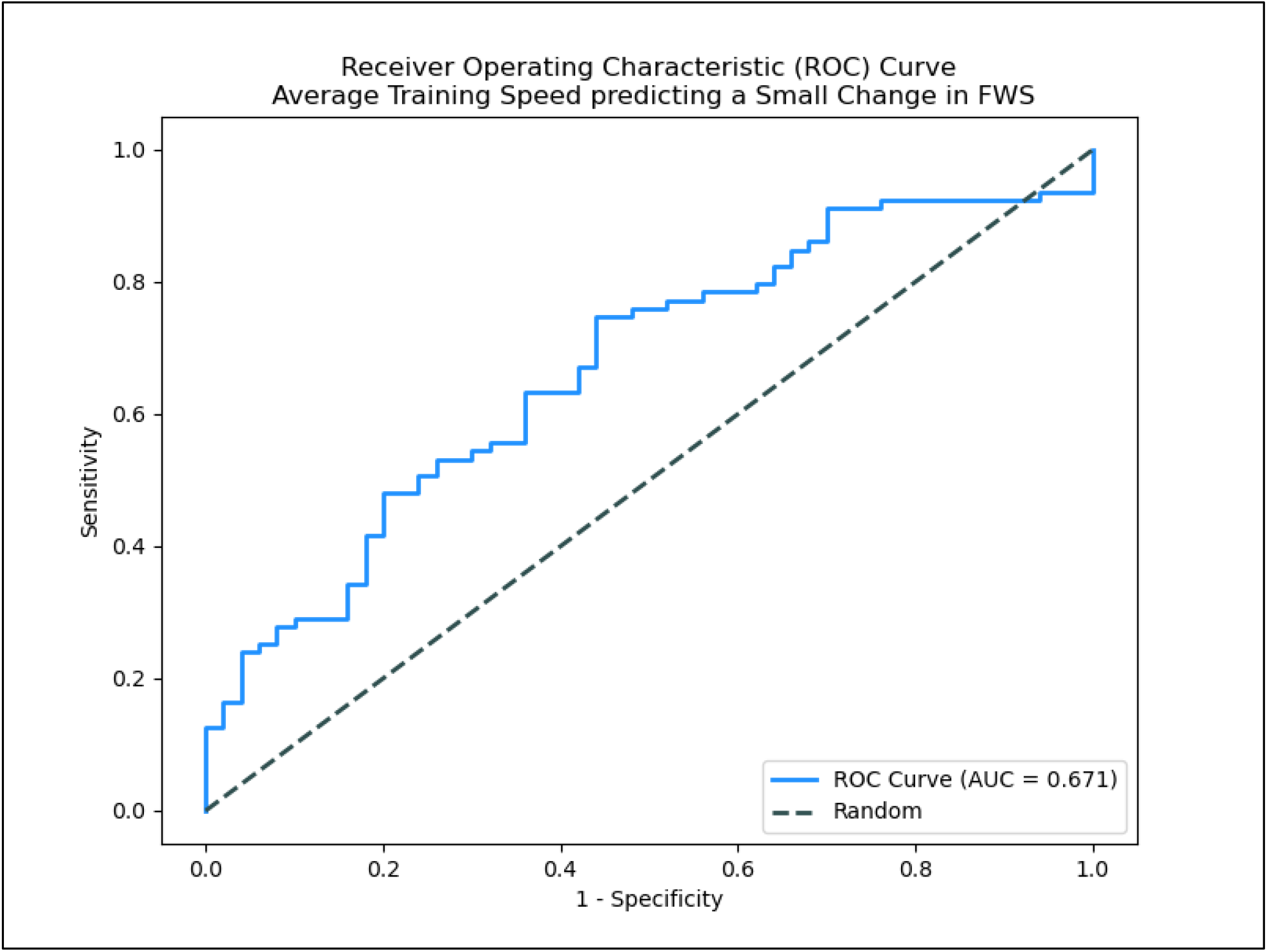
*FWS = Fastest Walking Speed; small change = a pre-to-post change in FWS distance of ≥ 0*.*1 meters/second*.

**Supplemental Figure 2.**
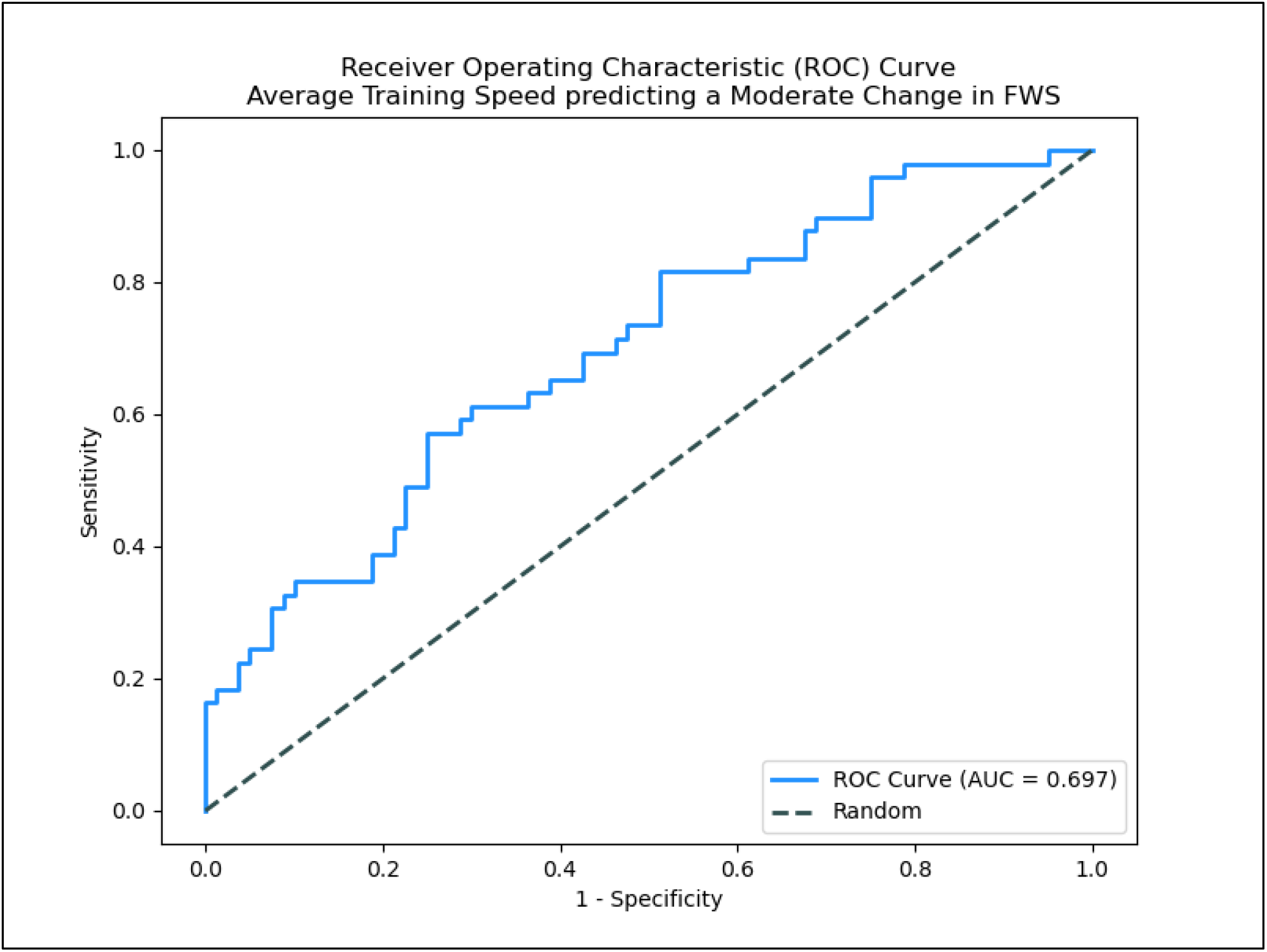
*FWS = Fastest Walking Speed; moderate change = a pre-to-post change in FWS distance of ≥ 0*.*2 meters/second*.

