## Supplemental Material for "Using Exercise Intensity to Predict a Minimal Clinically Important Difference in the Six-Minute Walk Test in People with Chronic Stroke"

*Appendix*

Additional Analyses

Two separate receiver operating characteristic (ROC) curves were used to analyze if exercise training speed is predictive of individuals attaining a (1) small or (2) moderate change in the FWS.[34] A small FWS change was defined as a change of 0.1m/s with a moderate change defined as a change of 0.2m/s.

Results

*Small Change in FWS*

Seventy-nine (61.2%) participants had a change of at least 0.1m/s in FWS. Average training speed had poor, significant accuracy (AUC [95% CI] = 0.671 [0.577 - 0.765], $p < 0.001$) of predicting a small change in FWS (Supplementary Figure 1).

*Moderate Change in FWS*

Forty-nine (38.3%) participants had a change of at least 0.2m/s in FWS. Average training speed had poor, significant accuracy (AUC [95% CI] = 0.697 [0.605- 0.790], $p < 0.001$) of predicting a moderate change in FWS (Supplementary Figure 2).

**Supplemental Figure 1.**

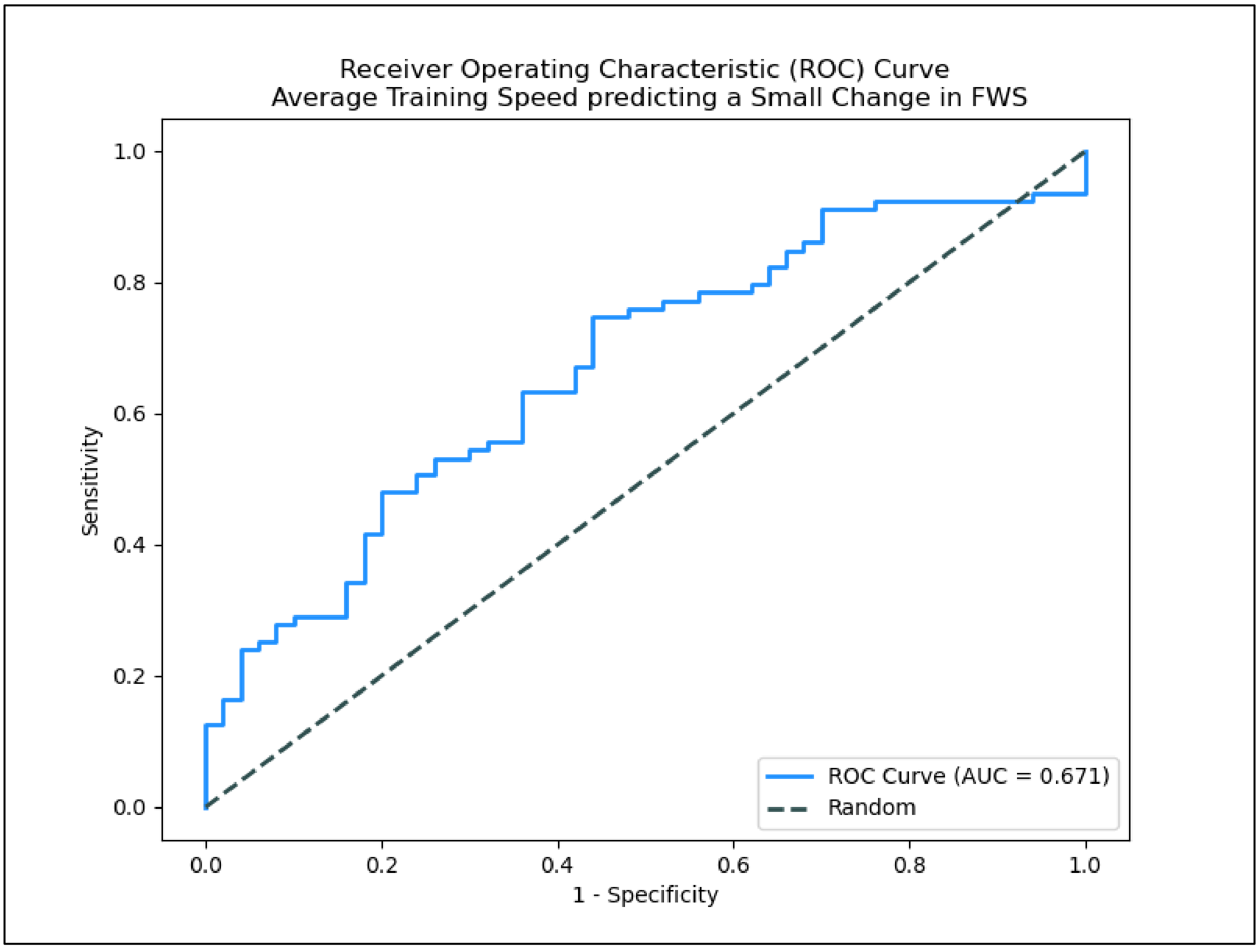


*FWS = Fastest Walking Speed;*
*small change = a pre-to-post change in FWS distance of* $\geq$ *0.1 meters/second.*

**Supplemental Figure 2.**

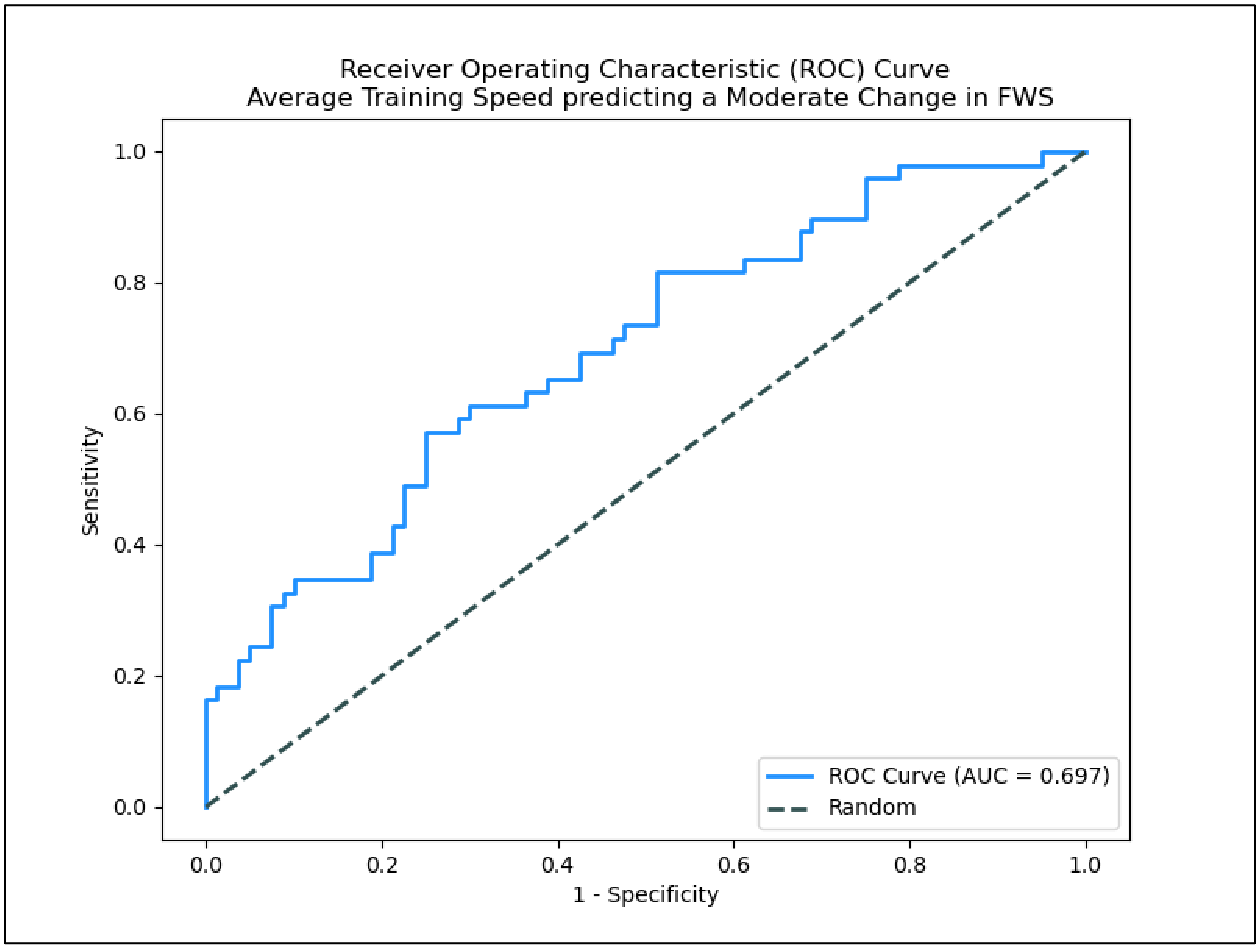


*FWS = Fastest Walking Speed;*
*moderate change = a pre-to-post change in FWS distance of ≥ 0.2 meters/second.*
